# Alcohol use and the pre-exposure prophylaxis continuum of care among men in rural South Africa

**DOI:** 10.1101/2024.03.23.24304770

**Authors:** Alison C. Castle, Jacob Busang, Jaco Dreyer, Carina Herbst, Nonhlanhla Okesola, Natsayi Chimbindi, Thembelihle Zuma, Jana Jarolimova, Judith A. Hahn, Christina Psaros, Sheela V. Shenoi, Maryam Shahmanesh, Mark J. Siedner

## Abstract

**Introduction:** Despite freely available HIV pre-exposure prophylaxis (PrEP), HIV incidence among young men in South Africa is high. There is conflicting evidence around the association between alcohol use behaviors and PrEP utilization. We explore the impact of alcohol use on PrEP initiation and continuation among South African men.

**Methods:** We performed a secondary analysis of data from a trial that included men aged 16-29 randomly selected from a demographic surveillance site in KwaZulu-Natal. All participants were referred to HIV and sexual health services, where those who were HIV negative and sexually active were offered oral PrEP. Alcohol consumption was assessed at monthly visits and categorized as: non-drinking (0), low/moderate risk drinking (1-5), and high/very high-risk drinking (6-12) based on AUDIT-C criteria. Primary outcomes were PrEP initiation and PrEP continuation defined as refilling prescriptions for ≥3 months. We fitted logistic regression models, adjusted for potential clinical and demographic confounders, to estimate relationships between PrEP initiation/continuation and reported alcohol use.

**Results:** Of the 325 men in the analytic cohort, the average age was 22.9 years (SD 3.6) and 131 (40%) had high/very high-risk alcohol consumption (AUDIT-C score ≥6). Men with the highest risk alcohol use also reported more frequent condomless sex (89%, vs 68% in no alcohol group). We found the greatest uptake of PrEP among the high/very high-risk alcohol group (46/131, 35%), followed by the low/moderate-risk group (17/53, 32%) and the no alcohol group (25/141, 18%). Those with high-risk alcohol use remained more likely to initiate PrEP compared to the no alcohol group in multivariable models adjusted for confounders (aOR 2.44 95% CI 1.29-4.60; p-value 0.006). Overall, only 30% (26/88) of men remained on PrEP at 3 months. Men with high/very high-risk drinking had similar PrEP continuation at 3 months compared to men who reported no alcohol use (aOR 1.02 95% CI: 0.28-3.86, p=0.98).

**Conclusions:** High-risk alcohol use is common among men in rural South Africa and associated with increased PrEP initiation. However, PrEP continuation was low overall, and similar across all levels of alcohol use. Hazardous alcohol use should not discourage PrEP implementation efforts to engage and retain young men.

## Introduction

South Africa continues to face one of the highest global burdens of HIV, with more than 8.45 million people living with the disease [1]. Notably, young men in the province of KwaZulu-Natal continue to experience the dual burden of HIV incidence and poor engagement with the HIV care cascade [2–5]. In attempts to mitigate transmission, various interventions such as male circumcision and treatment as prevention have been instituted [6, 7]. However, these methods have not led to the elimination of HIV in KwaZulu-Natal. This underscores the critical need for the development and implementation of more effective prevention strategies, including for young men.

Oral pre-exposure prophylaxis (PrEP) was approved in South Africa in 2016 and its implementation has seen over 880,000 individuals initiated on the regimen [8]. However, most PrEP implementation programs have focused on adolescent girls, young women, sex workers, and men who have sex with men, leaving a noticeable gap in access for heterosexual men [9–11]. Whereas men are now included in expanded PrEP access as part of the South African National Strategic Plan for HIV, TB, and STIs, their uptake of PrEP remains alarmingly low [12–15]. This gendered difference in PrEP uptake mirrors the broader gender disparity in healthcare engagement across Southern Africa [4, 16–18]. For example, in Eswatini men were nearly three times less likely to initiate PrEP than women [15]. Compared to women, men are less likely to engage in care, remain in care, and adhere to medical recommendations, with few men taking HIV preventive healthcare measures like PrEP [19]. Among heterosexual men, reasons for PrEP refusal include misperceptions about HIV as a curable disease, underestimation of personal risk, apprehensions about pill adherence, and stigma [14, 15].

Central to the questions surrounding PrEP utilization is the role of alcohol consumption. Alcohol is frequently used with other risky behaviors, heightening HIV transmission by impairing judgment and leading to unprotected intercourse, multiple sexual partners, and sex while under the influence [20, 21]. In South Africa, alcohol consumption is widely reported, particularly among men: the 2018 Global Status Report ranked South Africa as having one of the most concerning patterns of alcohol consumption, with the nation leading in reported use within Africa [22]. More than 70% of men who reported alcohol use engaged in heavy episodic drinking [22]. In various contexts, alcohol use has been found to be both a facilitator and barrier to PrEP utilization [21]. While some research suggests that men report alcohol use and condomless sex as motivators for seeking PrEP initiation [12, 21], others have found alcohol consumption might reduce the willingness to use PrEP [23]. In sub-Saharan Africa, female sex workers and those in serodiscordant relationships expressed concerns over the perceived toxic effects when mixing alcohol, often consumed before sexual activities, and PrEP [24]. Nonetheless, there is no evidence to suggest significant interactions between alcohol and PrEP [25]. Individuals who drink alcohol may represent crucial targets for PrEP due to their association with risky sexual behaviors. However, the relationship between high-risk alcohol consumption and PrEP dynamics among men in South Africa remains largely unexplored.

In this study, we leveraged data from a longitudinal HIV prevention study in rural KwaZulu-Natal, South Africa to assess the interplay between alcohol consumption, PrEP initiation, and PrEP continuation among young men. We hypothesized that individuals who report high-risk alcohol use may be more likely to initiate PrEP, but less likely to continue the preventative therapy compared to individuals who report no or low risk alcohol use.

## Methods

### Study population and procedures

We analyzed data from the Isisekelo Sempilo study, which was an open-label 2×2 randomized factorial trial that was conducted from March 2020-August 2022 in South Africa. The study evaluated the effectiveness of various referral mechanisms for linkage to care for HIV prevention services [26]. In brief, participants included men and women aged 16-29 years in Africa Health Research Institute’s Health and Demographic Surveillance Site (HDSS) in the uMkhanyakude district of KwaZulu-Natal [27]. The HDSS was used to randomly select 3000 participants, stratified by gender, from a primarily rural, economically disadvantaged, and high HIV prevalence region.

Potential participants were approached at their homes and invited to participate in the study. All enrolled participants, after being randomized to one of the four study arms, were provided with barcoded referral slips and an appointment to attend one of the youth friendly study clinics within the HDSS for HIV prevention and sexual reproductive health (SRH) services. The four arms of the study differed by referral pathways and included 1) the enhanced standard of care arm where participants attended one of the study clinics without additional referral pathways 2) the enhanced standard of care arm as above with peer navigator support [28] 3) home-based self-collection of samples for sexually transmitted infection (STI) testing with referral to study clinics to receive results and SRH services, and 4) combined peer navigator support and home-based STI specimen collection with referral to study clinics. All clinic attendees were offered HIV testing and SRH services, including education on PrEP. If participants were HIV seronegative, sexually active, and interested in oral PrEP, they were provided with one month’s supply of generic tenofovir disoproxil fumarate and emtricitabine (TDF/FTC). Participants also completed questionnaires on sociodemographic measures, alcohol consumption behaviors over the preceding 30 days, and sexual behaviors. Participants were requested to visit the study clinics at one month, followed by every 3 months for HIV re-testing, adherence counseling, safety monitoring, and PrEP refills while taking PrEP. Clinic attendance was recorded through scanned barcodes on referral slips. If participants didn’t have their slips, an algorithm utilizing demographic details was used to identify them.

### Study definitions

Our primary exposure of interest was self-reported alcohol use behavior. We ascertained alcohol use using a standardized questionnaire which queried whether 1) respondents had ever consumed an alcoholic drink in their life; 2) the number of days they had consumed alcohol over the last 30 days and 3) the number of days they had consumed 5 or more drinks over the last 30 days. We mapped the question responses to the validated Alcohol Use Disorders Identification Test-Consumption (AUDIT-C) questionnaire and categorized alcohol use as: no alcohol use (AUDIT-C score 0), low/moderate-risk alcohol use (AUDIT-C score 1–5), high/very high-risk alcohol use (AUDIT-C score 6-12).[29, 30] These scores align with prior literature showing increased risk for alcohol use disorder with increasing AUDIT-C scores [31]. For men with repeat AUDIT-C scores obtained at follow up visits, the maximum AUDIT-C scores were used for analyses.

Our primary outcomes of interest were 1) PrEP initiation and 2) PrEP continuation among those who initiated PrEP. Participants were offered oral PrEP at each clinic visit if they had negative HIV rapid testing and were considered at risk for HIV according to the South African screening guidelines [32]. Therefore, persons who declined HIV testing, had positive HIV testing, or who were not sexually active were excluded from our PrEP-eligible cohort. PrEP continuation was defined as receiving on time PrEP refill prescriptions from the study clinics at or beyond 3 months of the PrEP initiation date. We use the terms continuation and persistence synonymously, as defined by the World Health Organization [33]. As the highest drop off for PrEP usage is after one month [10, 34, 35], we extended our outcome to be 6 months in sensitivity analyses. Additionally, we evaluated the number of HIV seroconversions among all men in our cohort at the 12-month end-line study visit.

We considered smoking, drug use, sexual behavior, and socioeconomic status as potential confounders or mediators of the relationship between alcohol use and PrEP initiation or continuation in regression models. Participants who reported current or past smoking history were characterized as ever smokers. Reported drug use was defined as “none”, “cannabis”, or “club drugs/other”. Participants reported number of sex partners within the last year, ranging from 0, 1, 2-4, or >5 partners.. Sexual behaviors including 1) knowledge of sexual partner’s HIV status, 2) reported sex while using alcohol or drugs, and 3) recent condomless sex within 3 months were all defined as binary variables. Socioeconomic status was estimated using household asset ownership data collected within the HDSS annual surveys to generate a relative wealth index, as developed by Filmer and Pritchett [27, 36].

### Statistical Methods

For the purposes of our analyses, we included only men with completed alcohol data at baseline. We first described participants’ characteristics stratified by alcohol use categories and explored differences between groups using Kruskal-Wallis testing. To explore the associations between alcohol use and PrEP initiation among eligible men, we fitted logistic regression models, with and without adjustment for potential confounders. The following covariates were included: age, socioeconomic status, knowledge of partner’s HIV status, male circumcision, and number of clinic visits. Reported sex while using alcohol or drugs, recent condomless sex, and number of sexual partners were not included as covariates in primary models as these were considered to be on the causal pathway. Further, drug use and smoking demonstrated collinearity with alcohol use and were not included in the final models. We then conducted a survival analysis with the Kaplan-Meier estimator to compare the duration of PrEP use, stratified by alcohol use categories, and performed a log-rank test to compare differences in PrEP continuation. Participants were considered to have discontinued PrEP 30 days after their last filled prescription or if they reported a specific stop date. We then fitted logistic regression models with the outcomes of PrEP continuation at 3 months and 6 months, with and without adjustment for age, socioeconomic status, knowledge of partner’s HIV status, and male circumcision. Men were excluded from the primary analysis if they did not have complete alcohol data obtained at the study clinic visits. However, in sensitivity analyses, we included men with completed alcohol data obtained at the 12-month end-line study survey. Statistical analyses were conducted in Stata (Version 17, StataCorp, College Station, Texas, USA).

### Ethical considerations

The institutional review boards approved the Isisekelo Sempilo study at the University of KwaZulu-Natal Biomedical Research Ethics Committee (BREC/00000473/2019) and University College London Research Ethics Committee (5672/003). Written informed consent was obtained from participants aged 18-29 years; written assent was obtained from participants aged 16-17 years, with written consent from their parents or guardian.

## Results

### Sample characteristics

Of the randomly selected men within the catchment area (n=1,500), 847 men (56%) were enrolled in the Isisekelo Sempilo study (**Figure 1**), of whom 528 (62%) attended a study clinic at least once. We excluded 203 men due to missing alcohol data (n=107), positive HIV testing (n=20), declined HIV testing at every study visit (n=29), or report that they were not sexually active (n=47), leaving 325 men eligible for PrEP in the analytic cohort (**Figure 1**). The average age of participants was 22.9 years (SD 3.6), with older ages among high/very high-risk alcohol use groups (24.4 years versus 21.6 years in no alcohol group, p<0.001) (**Table 1**). Of the 325 men, 40% (141) had high/very high-risk alcohol consumption behaviors (AUDIT-C score ≥6). Socioeconomic status and male circumcision rates were comparable across the three alcohol use groups. Men with the highest risk alcohol use reported greater condomless sex (89.3%, vs 68.1% in no alcohol group, p-value<0.001), and having more than 5 sexual partners in the last year (16.0%, vs 5.0% in no alcohol group, p-value=0.003). Knowledge of sexual partner’s HIV status was reported by approximately one third of participants (total group 35.4%, n=115/325).

**Figure 1:**
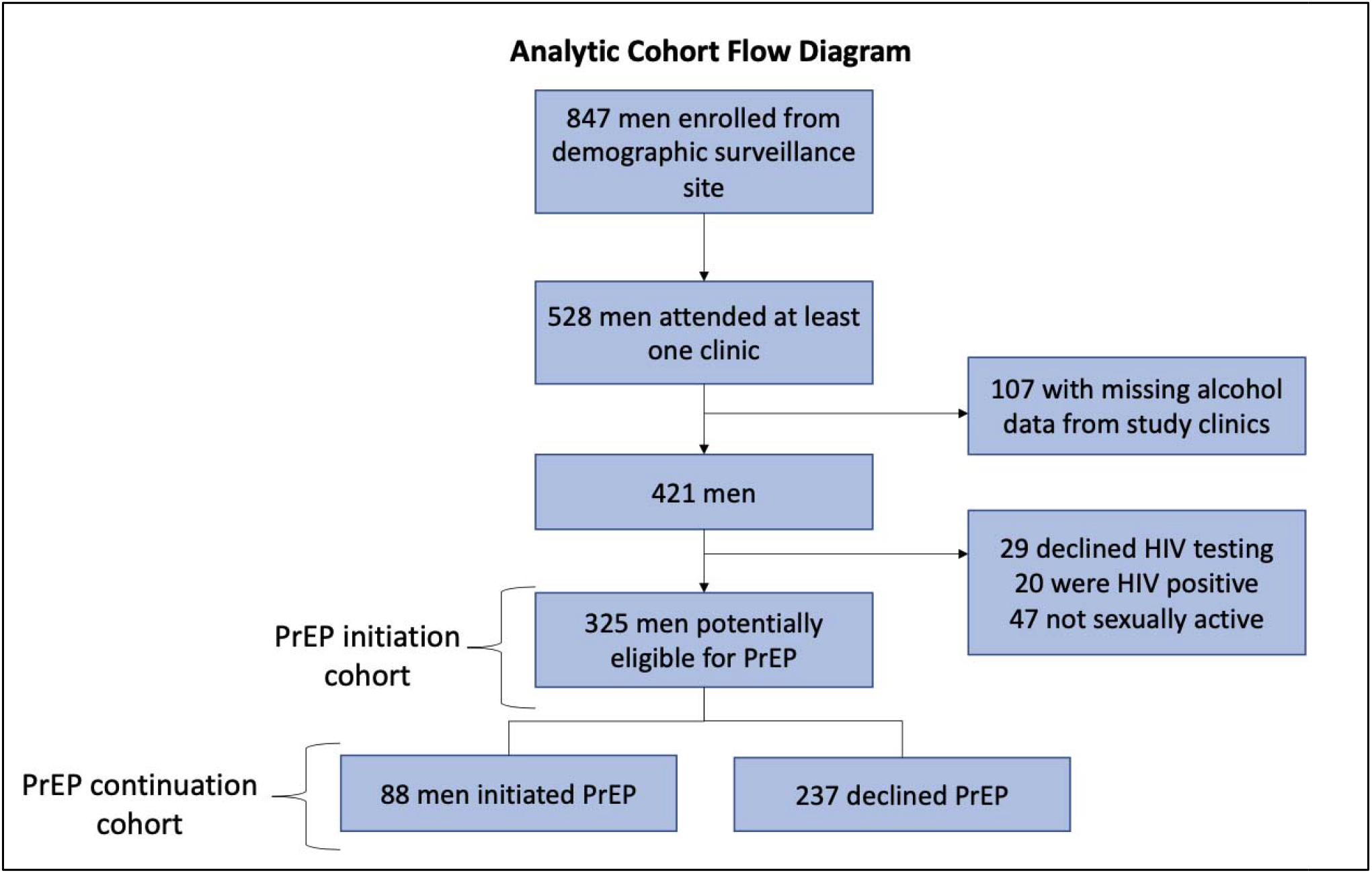
Analytic cohort.

**Table 1:** Characteristics of Men by Alcohol Consumption Groups.

**Figure 1: Analytic cohort**
| **Characteristic** | **Total**<br>**(N=325)** | **No alcohol**<br>**(N=141)** | **Low/moderate**<br>**risk alcohol**<br>**(N=53)** | **High/very high**<br>**risk alcohol**<br>**(N=131)** | **p-value<sup>^</sup>** |
| --- | --- | --- | --- | --- | --- |
| Age (years) | 22.9 (3.6) | 21.6 (3.1) | 22.6 (3.4) | 24.4 (3.5) | <0.001 |
| Socioeconomic status |  |  |  |  |  |
| Lowest | 28 (8.6) | 12 (8.5) | 4 (7.6) | 12 (9.2) | 0.366 |
| Low | 65 (20.0) | 29 (20.6) | 13 (24.5) | 23 (17.6) |  |
| Middle | 73 (22.5) | 39 (27.7) | 12 (22.6) | 22 (16.8) |  |
| High | 51 (15.7) | 19 (13.5) | 7 (13.2) | 25 (19.1) |  |
| Highest | 61 (18.8) | 27 (19.2) | 11 (20.8) | 23 (17.6) |  |
| Unknown | 47 (14.5) | 15 (10.6) | 6 (11.3) | 26 (19.9) |  |
| Ever smoker | 95 (29.2) | 16 (11.4) | 10 (18.9) | 69 (52.7) | <0.001 |
| Drug use |  |  |  |  |  |
| None | 266 (81.9) | 128 (90.8) | 44 (83.0) | 94 (71.8) | 0.002 |
| Cannabis | 56 (17.2) | 12 (8.5) | 8 (15.1) | 36 (27.5) |  |
| Club drugs/Other | 3 (0.9) | 1 (0.7) | 1 (1.9) | 1 (0.8) |  |
| Circumcised | 245 (75.4) | 105 (74.5) | 38 (71.7) | 102 (77.9) | 0.435 |
| Recent condomless sex | 255 (78.5) | 96 (68.1) | 42 (79.3) | 117 (89.3) | <0.001 |
| Sex while using alcohol<br>or drugs | 124 (38.2) | 11 (7.8) | 19 (35.9) | 94 (71.8) | <0.001 |
| Knowledge of sexual<br>partner's HIV status | 115 (35.4) | 48 (34.0) | 19 (35.9) | 48 (36.6) | 0.111 |
| Number sex partners in<br>last year |  |  |  |  |  |
| 0-1 partners | 125 (38.5) | 69 (48.9) | 16 (30.2) | 40 (30.5) | 0.003 |
| 2-4 partners | 139 (42.8) | 52 (36.9) | 24 (45.3) | 63 (48.1) |  |
| >5 partners | 34 (10.5) | 7 (5.0) | 6 (11.3) | 21 (16.0) |  |
| Declined to answer | 27 (8.3) | 13 (9.2) | 7 (13.2) | 7 (5.3) |  |

Values are mean (standard deviation) or number (%)
<sup>^</sup>p-values calculated for comparisons between alcohol consumption groups by Kruskal-Wallis testing

### Association between alcohol use and PrEP initiation among eligible men

We found the greatest uptake of PrEP among the high/very high-risk alcohol use group (46/131, 35%), followed by the low/moderate risk group (17/53, 32%) and the no alcohol group (25/141, 18%) (**Figure 2**). In univariate analyses, high/very high-risk alcohol use was positively associated with PrEP initiation compared to no alcohol use (OR 2.64 95%CI 1.50-4.65, p=0.001) (**Table 2**). In multivariable models adjusted for demographic and clinical confounders, the high/very high-risk alcohol group remained significantly more likely to initiate PrEP compared to the no alcohol group (aOR 2.44 95% CI 1.29-4.60; p-value 0.006). Men in the low/moderate alcohol use group also initiated PrEP more frequently than men who do not drink alcohol (aOR 2.20 95% CI 1.03-4.71, p=0.041). In sensitivity analyses, we included men with completed alcohol data obtained at the 12-month end-line survey. Therefore, 75 of the 107 men initially excluded were part of our sensitivity analyses; we found similar relationships persisted between alcohol use and PrEP initiation (Supplemental Figure 1, Supplemental Table 1).

**Figure 2:**
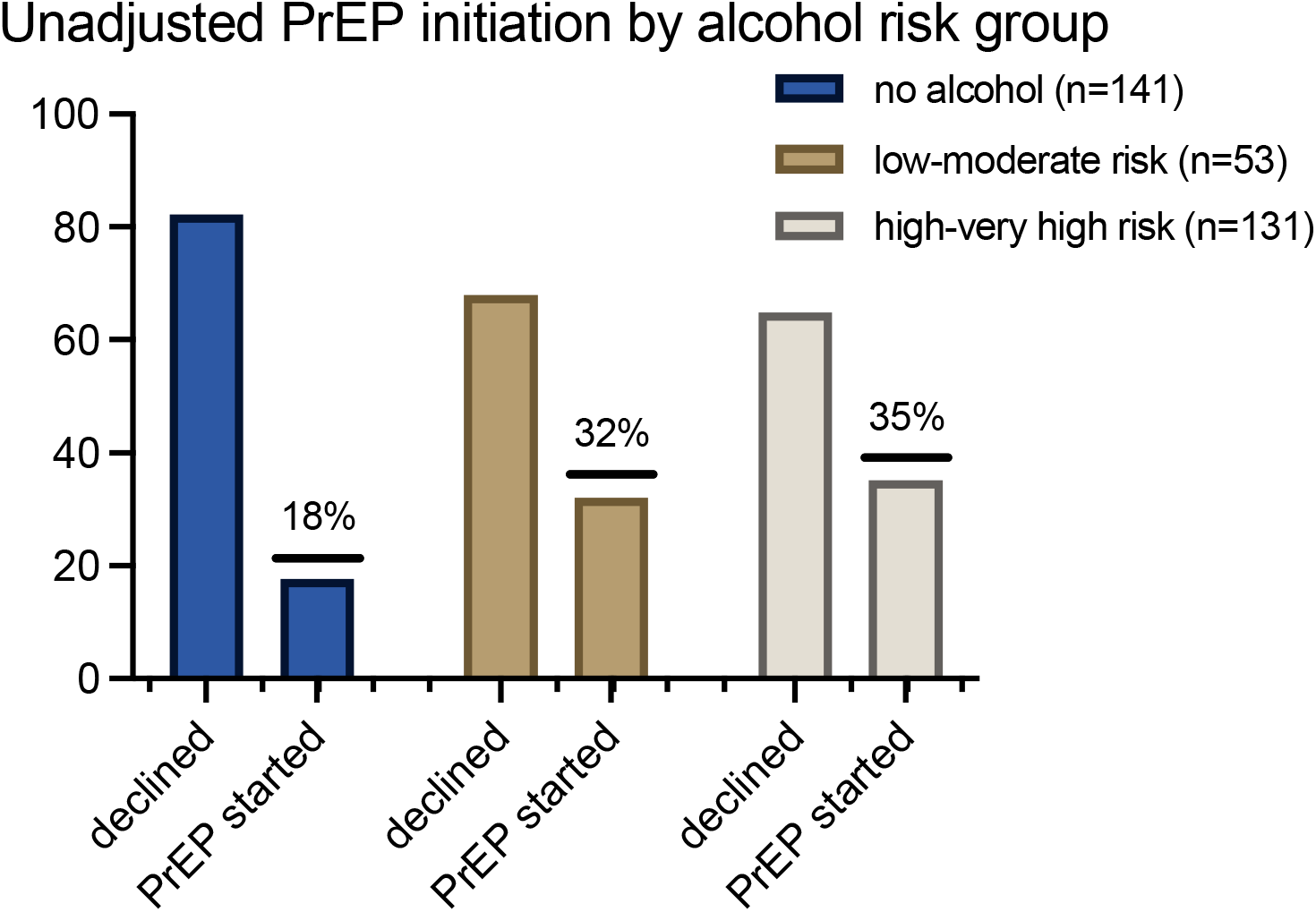
PrEP initiation among men stratified by alcohol risk group.

**Table 2:** Logistic regression models for PrEP initiation and PrEP continuation.

| Characteristic | Unadjusted Odds Ratio (95%CI) | p-value | Adjusted Odds Ratio (95% CI) | p-value |
| --- | --- | --- | --- | --- |
| **Model: PrEP initiation (n=325)** |  |  |  |  |
| No alcohol | Reference group |  |  |  |
| Low/moderate risk alcohol | 2.30 (1.11-4.75) | 0.024 | 2.20 (1.03-4.71) | 0.041 |
| High/very high risk alcohol | 2.64 (1.50-4.65) | 0.001 | 2.44 (1.29-4.60) | 0.006 |
| Age (years) | 1.04 (0.97-1.12) | 0.236 | 0.98 (0.90-1.06) | 0.586 |
| Socioeconomic status | 0.98 (0.85-1.12) | 0.745 | 0.96 (0.83-1.11) | 0.587 |
| Circumcised | 1.52 (0.95-2.43) | 0.083 | 1.16 (0.70-1.95) | 0.565 |
| Knowledge of sexual partner's HIV status | 2.09 (1.30-3.34) | 0.002 | 1.65 (0.95-2.85) | 0.074 |
| Number of visits | 1.45 (1.22-1.72) | <0.001 | 1.28 (1.06-1.55) | 0.011 |
| **Model: PrEP continuation at 3 months (n=88)** |  |  |  |  |
| No alcohol | Reference group |  |  |  |
| Low/moderate risk alcohol | 0.68 (0.14-3.19) | 0.624 | 0.41 (0.07-2.21) | 0.298 |
| High/very high risk alcohol | 1.86 (0.62-5.55) | 0.269 | 1.02 (0.28-3.86) | 0.980 |
| Age (years) | 1.13 (0.98-1.30) | 0.087 | 1.11 (0.94-1.33) | 0.221 |
| Socioeconomic status | 1.16 (0.90-1.50) | 0.260 | 1.16 (0.87-1.55) | 0.323 |
| Circumcised | 3.64 (0.82-16.16) | 0.089 | 4.86 (0.94-25.16) | 0.060 |
| Knowledge of sexual partner's HIV status | 3.80 (1.44-10.0) | 0.007 | 3.03 (1.10-8.34) | 0.032 |
| **Model: PrEP continuation at 6 months n=88** |  |  |  |  |
| No alcohol | Reference group |  |  |  |
| Low/moderate risk alcohol | 0.70 (0.11-4.32) | 0.701 | 0.48 (0.07-3.32) | 0.459 |
| High/very high risk alcohol | 1.85 (0.53-6.50) | 0.336 | 1.14 (0.26-5.09) | 0.859 |
| Age (years) | 1.11 (0.95-1.30) | 0.174 | 1.07 (0.89-1.29) | 0.470 |
| Socioeconomic status | 1.16 (0.87-1.56) | 0.320 | 1.14 (0.83-1.56) | 0.418 |
| Circumcised | 2.19 (0.50-9.6) | 0.296 | 2.42 (0.49-12.04) | 0.279 |
| Knowledge of sexual partner's HIV status | 3.87 (1.26-11.94) | 0.018 | 3.09 (0.97-9.88) | 0.057 |

### Association between alcohol use and PrEP continuation among men who initiated PrEP

Out of the 88 participants who initiated PrEP, 83 discontinued PrEP within the 12-month study observation period. The median duration of PrEP use was 30 days for the no alcohol group, 44 days for the low/moderate risk alcohol group, and 53 days for the high/very high-risk alcohol group (**Figure 3**, **Table 3**, log rank test p-value=0.570).

**Figure 3:**
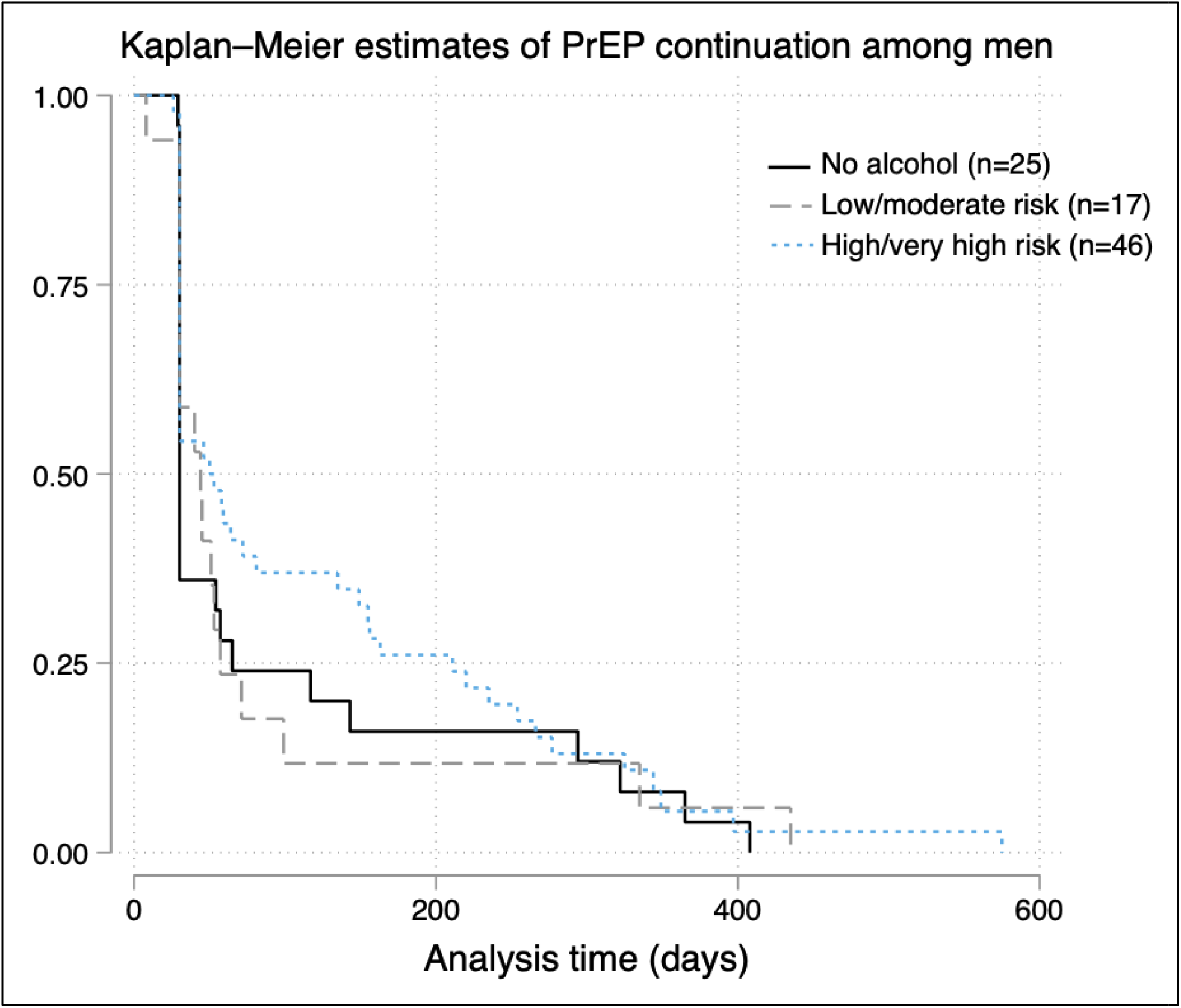
Kaplan-Meier analysis of PrEP continuation among men. The Kaplan-Meier estimates of PrEP continuation among men illustrate the survival probabilities across the three alcohol groups. The median PrEP continuation time was 30 days in the no alcohol group, 44 days in the low/moderate-risk alcohol group, and 53 days in the high/very high-risk group. However, there is no statistical difference in PrEP continuation rates over time among these groups as indicated by a log-rank p-value of 0.501.

**Table 3:** Kaplan-Meier survival estimates for PrEP continuation among men.

| Time<br>(days) | At<br>Risk | Fail | Net<br>Lost | Survivor<br>Function | Std. error | 95% Confidence<br>Interval |  |
| --- | --- | --- | --- | --- | --- | --- | --- |
| No alcohol group |  |  |  |  |  |  |  |
| 29 | 25 | 1 | 0 | 0.9600 | 0.0392 | 0.7484 | 0.9943 |
| **30\*** | **24** | **15** | **0** | **0.3600** | **0.096** | **0.1819** | **0.542** |
| 54 | 9 | 1 | 0 | 0.3200 | 0.0933 | 0.1524 | 0.5015 |
| 57 | 8 | 1 | 0 | 0.2800 | 0.0898 | 0.1242 | 0.4598 |
| 65 | 7 | 1 | 0 | 0.2400 | 0.0854 | 0.0976 | 0.4167 |
| 117 | 6 | 1 | 0 | 0.2000 | 0.08 | 0.0728 | 0.372 |
| 143 | 5 | 1 | 0 | 0.1600 | 0.0733 | 0.0502 | 0.3254 |
| 294 | 4 | 1 | 0 | 0.1200 | 0.065 | 0.0303 | 0.2766 |
| 322 | 3 | 1 | 0 | 0.0800 | 0.0543 | 0.0139 | 0.2249 |
| 365 | 2 | 1 | 0 | 0.0400 | 0.0392 | 0.0029 | 0.1699 |
| 408 | 1 | 1 | 0 | 0 | . | . | . |
| Low/moderate-risk alcohol group |  |  |  |  |  |  |  |
| 8 | 17 | 1 | 0 | 0.9412 | 0.0571 | 0.6502 | 0.9915 |
| 30 | 16 | 6 | 0 | 0.5882 | 0.1194 | 0.3254 | 0.7782 |
| 40 | 10 | 1 | 0 | 0.5294 | 0.1211 | 0.2762 | 0.7303 |
| **44\*** | **9** | **1** | **0** | **0.4706** | **0.1211** | **0.2296** | **0.6797** |
| 45 | 8 | 1 | 0 | 0.4118 | 0.1194 | 0.1858 | 0.6264 |
| 51 | 7 | 1 | 0 | 0.3529 | 0.1159 | 0.1448 | 0.5704 |
| 53 | 6 | 1 | 0 | 0.2941 | 0.1105 | 0.1071 | 0.5115 |
| 57 | 5 | 1 | 0 | 0.2353 | 0.1029 | 0.0731 | 0.4492 |
| 71 | 4 | 1 | 0 | 0.1765 | 0.0925 | 0.0435 | 0.383 |
| 99 | 3 | 1 | 0 | 0.1176 | 0.0781 | 0.0196 | 0.312 |
| 335 | 2 | 1 | 0 | 0.0588 | 0.0571 | 0.0039 | 0.235 |
| 435 | 1 | 1 | 0 | 0 | . | . | . |
| High/very high-risk alcohol group |  |  |  |  |  |  |  |
| 26 | 46 | 1 | 0 | 0.9783 | 0.0215 | 0.8555 | 0.9969 |
| 30 | 45 | 20 | 0 | 0.5435 | 0.0734 | 0.3901 | 0.6737 |
| 46 | 25 | 1 | 0 | 0.5217 | 0.0737 | 0.3696 | 0.6536 |
| 50 | 24 | 1 | 0 | 0.5 | 0.0737 | 0.3493 | 0.6333 |
| **53\*** | **23** | **1** | **0** | **0.4783** | **0.0737** | **0.3294** | **0.6127** |
| 58 | 22 | 1 | 0 | 0.4565 | 0.0734 | 0.3097 | 0.5919 |
| 59 | 21 | 1 | 0 | 0.4348 | 0.0731 | 0.2902 | 0.5708 |
| 64 | 20 | 1 | 0 | 0.413 | 0.0726 | 0.2711 | 0.5494 |
| 72 | 19 | 1 | 0 | 0.3913 | 0.072 | 0.2522 | 0.5278 |
| 81 | 18 | 1 | 0 | 0.3696 | 0.0712 | 0.2335 | 0.506 |
| 135 | 17 | 1 | 0 | 0.3478 | 0.0702 | 0.2152 | 0.4838 |
| 149 | 16 | 1 | 0 | 0.3261 | 0.0691 | 0.1972 | 0.4614 |
| 155 | 15 | 1 | 0 | 0.3043 | 0.0678 | 0.1795 | 0.4387 |
| 156 | 14 | 1 | 0 | 0.2826 | 0.0664 | 0.1622 | 0.4157 |
| 163 | 13 | 1 | 0 | 0.2609 | 0.0647 | 0.1452 | 0.3923 |
| 211 | 12 | 1 | 0 | 0.2391 | 0.0629 | 0.1286 | 0.3686 |
| 220 | 11 | 1 | 0 | 0.2174 | 0.0608 | 0.1124 | 0.3446 |
| 235 | 10 | 1 | 0 | 0.1957 | 0.0585 | 0.0967 | 0.3201 |
| 254 | 9 | 1 | 0 | 0.1739 | 0.0559 | 0.0815 | 0.2951 |
| 266 | 8 | 1 | 0 | 0.1522 | 0.053 | 0.0669 | 0.2697 |
| 277 | 7 | 1 | 0 | 0.1304 | 0.0497 | 0.053 | 0.2436 |
| 325 | 6 | 1 | 0 | 0.1087 | 0.0459 | 0.0399 | 0.2169 |
| 329 | 5 | 0 | 1 | 0.1087 | 0.0459 | 0.0399 | 0.2169 |
| 344 | 4 | 1 | 0 | 0.0815 | 0.0417 | 0.0238 | 0.1863 |
| 349 | 3 | 1 | 0 | 0.0543 | 0.0356 | 0.0108 | 0.1534 |
| 397 | 2 | 1 | 0 | 0.0272 | 0.0262 | 0.0023 | 0.1182 |
| 575 | 1 | 1 | 0 | 0 | . | . | . |

**Bolded\*** is the median survival time until PrEP discontinuation within each alcohol consumption group. Underline indicates the end of the 12-month study period for PrEP refills.

Twenty-four percent (6/25) of the no alcohol group, 18% (3/17) of the low/moderate risk group, and 37% (17/46) of the high/very high-risk group remained on PrEP at 3 months, respectively. In multivariable models, we found no significant association between the alcohol use groups and 3-month PrEP continuation when compared to the no alcohol group (high/very high-risk group aOR 1.02 95% CI: 0.28-3.86, p=0.980; low/moderate risk group aOR of 0.41 95% CI: 0.07-2.21, p=0.298) (**Table 2**). By the 6-month time point, 18 of the 88 participants (20%) remained on PrEP, with no differences in continuation based on alcohol consumption (**Table 2**).

Three men seroconverted during the 12-month study period. Among these, two reported high/very high-risk alcohol use behaviors, while the third reported no alcohol use. Notably one of the individuals in the high/very high-risk alcohol group denied sexual activity and therefore was not encouraged to take oral PrEP. The individual with no alcohol use declined PrEP. The third participant did initiate PrEP on his second clinic visit, however, did not return for follow up despite attempts made by the study team. He was found to have seroconverted at the 12-month end-line study assessment.

## Discussion

In one of the first studies examining alcohol and PrEP use in southern Africa, we found men reporting high/very high-risk alcohol use were significantly more likely to initiate PrEP than men who do not drink alcohol. This observation echoes findings from other studies suggesting that men engaging in riskier behaviors, such as hazardous alcohol consumption, perceive themselves to be at higher risk of HIV acquisition and therefore may be more inclined to start PrEP as a preventive measure [12, 14]. However, irrespective of alcohol use, we observed a sharp decline in PrEP continuation rates – a trend observed in other studies noting substantial PrEP discontinuation within a month of initiating treatment [10, 31, 34]. Reassuringly, men with the highest risk alcohol use showed similar continuation with oral PrEP as those who do not consume alcohol. Our results highlight the relationship between alcohol use, perceived HIV risk, and PrEP utilization, demonstrating that high risk alcohol use should not be a perceived barrier to PrEP provision in rural South Africa. On the contrary, given their reporting of high-risk sexual activities, it’s essential to prioritize engaging men, especially those who consume alcohol, in safe sexual practices and HIV prevention methods to curb the HIV incidence in the region.

Hazardous alcohol use has been hypothesized to affect every aspect of the PrEP care continuum, however, to date, the evidence, largely from the United States, has been mixed regarding alcohol use and PrEP initiation [21]. Qualitative studies have described alcohol use as a perceived deterrent for PrEP initiation, due to concerns about potential toxicity when it is used concurrently with alcohol [23, 24]. By contrast, our study among young South African men found alcohol use as a facilitator to initiating PrEP. These results are consistent with those of a study conducted in Buffalo City, South Africa in which men with heavy alcohol use were more likely to initiate the preventative therapy [12]. Such data imply that men who engage in high-risk alcohol use also recognize their associated risk of HIV acquisition, justifying the use of preventative therapy. Thus, strategies to engage these individuals should be amplified.

Evidence for the relationship between alcohol use and PrEP effective use and continuation is also mixed. Two prospective studies, including one in Uganda and Kenya among serodiscordant couples, observed a decrease in PrEP adherence and continuation with hazardous alcohol use [37, 38]. In contrast, recent studies indicate that those reporting alcohol consumption often show better adherence, supported by biomarkers confirming medication compliance [39, 40]. An illustrative example comes from a South African study involving pregnant and post-partum women where 1 in 3 women reported hazardous alcohol use. At 3 months, 58% of women were reportedly still using PrEP and alcohol use correlated with higher odds of PrEP continuation (aOR 1.54, 95% CI 1.16-2.06) [40]. In our study, while there was a higher inclination towards initiating PrEP among the high-risk alcohol group, continuation rates remained largely uniform irrespective of alcohol use. This aligns with a U.S. study among men who have sex with men, which found no differences in dried blood spot tenofovir diphosphate levels between those who used alcohol and club drugs and those who did not use substances [41]. In our study, although the median discontinuation time was marginally longer for the high/very high-risk alcohol group, it remained less than two months, indicating a general challenge in sustained PrEP usage. Thus our data suggest that the decision to initiate doesn’t translate to continued usage, and that barriers to PrEP maintenance, such as potential side effects, societal stigma, and lack of healthcare access, continue to exist for men, regardless of their alcohol consumption [21, 42].

Our results also shed light on the complex relationship between sexual behaviors and alcohol use. Men with the highest alcohol use reported frequent condomless sex and multiple sexual partners, reflecting the intersectionality of risk behaviors. A holistic approach, which addresses these interlinked behaviors comprehensively rather than separately, might generate more effective intervention results. To this end, creating interventions that directly address the barriers men face regarding preventative therapy could be pivotal for enhancing PrEP uptake and sustained use. For example, a community-based PrEP delivery program, which engaged men for PrEP at local bars in rural South Africa, demonstrated potential feasibility of this approach to engage a traditionally hard-to-reach demographic of young men [43]. Of the 136 men deemed eligible for PrEP outside of local bars, 27% completed clinical evaluation and initiated PrEP [43]. While a 27% initiation rate of PrEP is low, it is consistent with the PrEP initiation rate in our cohort and falls within the range of other prospective cohorts conducted across southern Africa [44–47].

### Strengths and Limitations

Our study has several strengths. We used a longitudinal study design, targeting young men without bias towards the gender of their sexual partners, and drew participants with a community-based engagement approach randomly selected from a Health and Demographic Surveillance Site in rural South Africa [27]. There were also important limitations. Firstly, PrEP continuation was defined by the act of refilling PrEP prescriptions at the study clinics, thereby lacking comprehensive data about how men were taking PrEP. For future studies, measuring tenofovir drug levels could offer more nuanced information regarding effective usage of the medication. Additionally, a substantial proportion of the cohort had missing alcohol data at the baseline study visit. This limitation was addressed by leveraging the endline AUDIT-C scores obtained on 70% of the men initially excluded in sensitivity analyses. Further, the AUDIT-C questions deviated from the standard approach, capturing monthly alcohol consumption rather than the conventional 12-month timeframe. Our study, while one of the first in southern Africa to focus on alcohol use and PrEP utilization, was small, particularly for measuring differences in PrEP continuation. The self-reported nature of the alcohol consumption and sexual behavior data might introduce recall and social desirability bias [48, 49]. Moreover, our findings from the rural KwaZulu-Natal context may not be generalizable to other regions or countries with different cultural and healthcare landscapes. Lastly, although our focus was on alcohol use, other factors such as mental health, interventions tailored to men’s sexual health, and access to peer support can also influence PrEP initiation and continuation and should be explored in future studies.

### Conclusions

Understanding the relationship between alcohol use and PrEP utilization is essential for creating effective interventions, particularly in the context of South Africa where heavy alcohol use and HIV transmission remain high. We found that men with high-risk alcohol use were more likely to initiate PrEP, but that PrEP continuation was universally low across all alcohol use levels. Our data suggest that offering PrEP to at-risk men who use alcohol is feasible, but that current PrEP delivery models are not achieving high levels of PrEP utilization in any groups of men. Rather, addressing PrEP continuation may require a multifaceted strategy with supportive, stigma-free environments, and minimal barriers to accessing prescriptions so all can access and sustain HIV preventive care.

## Competing interests

Dr. Hahn served as a consultant for Pear Therapeutics in 2022. All other authors declare no competing interests.

## Authors’ contributions

AC contributed to the conception and design of the study, performed the data analysis and interpretation, and drafted the manuscript. JB, MJS, and JD contributed to the data analyzation and interpretation. MS designed and conducted the study, secured funding, and helped to critically revise the manuscript for important intellectual content. JB, JD, CH, NO, NC, and TZ all contributed toward the design and collection of the data. All authors critically reviewed the manuscript and approved the final version.

## Acknowledgements

The authors would like to thank the residents of the Africa Health Research Institute’s Health and Demographic surveillance site who participated in the Isisekelo Sempilo study.

## Funding

This research was supported by the Fogarty International Center (D43 TW010543), the National Institute of Mental Health (R01MH114560), the National Institute of Allergy and Infectious Diseases (T32 AI007433), the National Heart, Lung, Blood Institute (K24 HL166024), and the National Institute on Alcohol Abuse and Alcoholism (K24 AA022586) of the National Institutes of Health. This research was also funded in part, by Wellcome (Grant number Wellcome Strategic Core award: 201433/Z/16/A). For the purpose of open access, the author has applied a CC BY public copyright license to any Author Accepted Manuscript version arising from this submission. The contents of this manuscript are solely the responsibility of the authors and do not necessarily represent the official views of the funders.

## Data Availability Statement

Data and the data dictionary defining each field can be accessed at https://data.ahri.org/index.php/catalog/1039 via the Africa Health Research Institute Data Repository. Please. Access can be granted after publication and upon approval of the proposed analyses by the Isisekelo Sempilo PI and completion of a data access agreement.

## Notes

### Clinical Protocols

https://bmcpublichealth.biomedcentral.com/articles/10.1186/s12889-022-12796-8#:~:text=07%20March%202022-,Isisekelo%20Sempilo%20study%20protocol%20for%20the%20effectiveness%20of%20HIV%20prevention,2%20factorial%20randomised%20controlled%20trial

### Author Declarations

The institutional review boards approved the Isisekelo Sempilo study at the University of KwaZulu-Natal Biomedical Research Ethics Committee (BREC/00000473/2019) and University College London Research Ethics Committee (5672/003).

## References

1. Africa SS. Mid-year population estimates, 2022. 2022:37.

2. Zuma K, Simbayi L, Zungu N, Moyo S, Marinda E, Jooste S, et al. The HIV Epidemic in South Africa: Key Findings from 2017 National Population-Based Survey. Int J Environ Res Public Health. 2022;19(13). Epub 20220701. doi: 10.3390/ijerph19138125. PubMed PMID: 35805784; PubMed Central PMCID: PMCPMC9265818.

3. Birdthistle I, Tanton C, Tomita A, de Graaf K, Schaffnit SB, Tanser F, et al. Recent levels and trends in HIV incidence rates among adolescent girls and young women in ten high-prevalence African countries: a systematic review and meta-analysis. Lancet Glob Health. 2019;7(11):e1521–e40. doi: 10.1016/S2214-109X(19)30410-3. PubMed PMID: 31607465; PubMed Central PMCID: PMCPMC7025003.

4. Baisley K, Chimbindi N, Mthiyane N, Floyd S, McGrath N, Pillay D, et al. High HIV incidence and low uptake of HIV prevention services: The context of risk for young male adults prior to DREAMS in rural KwaZulu-Natal, South Africa. PLoS One. 2018;13(12):e0208689. Epub 20181226. doi: 10.1371/journal.pone.0208689. PubMed PMID: 30586376; PubMed Central PMCID: PMCPMC6306176.

5. Akullian A, Vandormael A, Miller JC, Bershteyn A, Wenger E, Cuadros D, et al. Large age shifts in HIV-1 incidence patterns in KwaZulu-Natal, South Africa. Proc Natl Acad Sci U S A. 2021;118(28). Epub 2021/07/11. doi: 10.1073/pnas.2013164118. PubMed PMID: 34244424; PubMed Central PMCID: PMCPMC8285891.

6. Tanser F, Kim HY, Vandormael A, Iwuji C, Barnighausen T. Opportunities and Challenges in HIV Treatment as Prevention Research: Results from the ANRS 12249 Cluster-Randomized Trial and Associated Population Cohort. Curr HIV/AIDS Rep. 2020;17(2):97–108. doi: 10.1007/s11904-020-00487-1. PubMed PMID: 32072468; PubMed Central PMCID: PMCPMC7072051.

7. Rosenberg MS, Gomez-Olive FX, Rohr JK, Kahn K, Barnighausen TW. Are circumcised men safer sex partners? Findings from the HAALSI cohort in rural South Africa. PLoS One. 2018;13(8):e0201445. Epub 20180801. doi: 10.1371/journal.pone.0201445. PubMed PMID: 30067842; PubMed Central PMCID: PMCPMC6070310.

8. Watch P. The Global PrEP Watcher: Country Overview South Africa. 2023.

9. Beesham I, Joseph Davey DL, Beksinska M, Bosman S, Smit J, Mansoor LE. Daily Oral Pre-exposure Prophylaxis (PrEP) Continuation Among Women from Durban, South Africa, Who Initiated PrEP as Standard of Care for HIV Prevention in a Clinical Trial. AIDS Behav. 2022;26(8):2623–31. Epub 20220205. doi: 10.1007/s10461-022-03592-x. PubMed PMID: 35122575; PubMed Central PMCID: PMCPMC9252967.

10. Celum CL, Bukusi EA, Bekker LG, Delany-Moretlwe S, Kidoguchi L, Omollo V, et al. PrEP use and HIV seroconversion rates in adolescent girls and young women from Kenya and South Africa: the POWER demonstration project. J Int AIDS Soc. 2022;25(7):e25962. doi: 10.1002/jia2.25962. PubMed PMID: 35822945; PubMed Central PMCID: PMCPMC9278271.

11. Medina-Marino A, Bezuidenhout D, Ngwepe P, Bezuidenhout C, Facente SN, Mabandla S, et al. Acceptability and feasibility of leveraging community-based HIV counselling and testing platforms for same-day oral PrEP initiation among adolescent girls and young women in Eastern Cape, South Africa. J Int AIDS Soc. 2022;25(7):e25968. doi: 10.1002/jia2.25968. PubMed PMID: 35872602; PubMed Central PMCID: PMCPMC9309460.

12. Smith PJ, Daniels J, Bekker LG, Medina-Marino A. What motivated men to start PrEP? A cross-section of men starting PrEP in Buffalo city municipality, South Africa. BMC Public Health. 2023;23(1):418. Epub 20230302. doi: 10.1186/s12889-023-15306-6. PubMed PMID: 36864381; PubMed Central PMCID: PMCPMC9979577.

13. Vogelzang M, Terris-Prestholt F, Vickerman P, Delany-Moretlwe S, Travill D, Quaife M. Cost-Effectiveness of HIV Pre-exposure Prophylaxis Among Heterosexual Men in South Africa: A Cost-Utility Modeling Analysis. J Acquir Immune Defic Syndr. 2020;84(2):173–81. doi: 10.1097/QAI.0000000000002327. PubMed PMID: 32141959.

14. Hannaford A, Lim J, Moll AP, Khoza B, Shenoi SV. ’PrEP should be for men only’: Young heterosexual men’s views on PrEP in rural South Africa. Glob Public Health. 2020;15(9):1337–48. Epub 20200324. doi: 10.1080/17441692.2020.1744680. PubMed PMID: 32207661.

15. Berner-Rodoreda A, Geldsetzer P, Barnighausen K, Hettema A, Barnighausen T, Matse S, et al. “It’s hard for us men to go to the clinic. We naturally have a fear of hospitals.” Men’s risk perceptions, experiences and program preferences for PrEP: A mixed methods study in Eswatini. PLoS One. 2020;15(9):e0237427. Epub 20200923. doi: 10.1371/journal.pone.0237427. PubMed PMID: 32966307; PubMed Central PMCID: PMCPMC7510987.

16. UNAIDS. Male engagement in HIV testing, treatment, and prevention in eastern and southern Africa —A framework for action. 2022.

17. Govender E, Abdool Karim Q. Understanding women and men’s acceptability of current and new HIV prevention technologies in KwaZulu-Natal, South Africa. AIDS Care. 2018;30(10):1311–4. Epub 20180618. doi: 10.1080/09540121.2018.1488027. PubMed PMID: 29914270.

18. Randera-Rees S, Clarence Safari W, Gareta D, Herbst K, Baisley K, Grant AD. Can we find the missing men in clinics? Clinic attendance by sex and HIV status in rural South Africa. Wellcome Open Res. 2021;6:169. Epub 20230821. doi: 10.12688/wellcomeopenres.16702.2. PubMed PMID: 37767058; PubMed Central PMCID: PMCPMC10521066.

19. UNAIDS. Joint United Nations Programme on HIV/AIDS. The blind spot: reaching out to men and boys. 2018.

20. Luehring-Jones P, Palfai TP, Tahaney KD, Maisto SA, Simons J. Pre-Exposure Prophylaxis (PrEP) Use is Associated With Health Risk Behaviors Among Moderate- and Heavy-Drinking MSM. AIDS Educ Prev. 2019;31(5):452–62. doi: 10.1521/aeap.2019.31.5.452. PubMed PMID: 31550196.

21. Oldfield BJ, Edelman EJ. Addressing Unhealthy Alcohol Use and the HIV Pre-exposure Prophylaxis Care Continuum in Primary Care: A Scoping Review. AIDS Behav. 2021;25(6):1777–89. Epub 20201120. doi: 10.1007/s10461-020-03107-6. PubMed PMID: 33219492; PubMed Central PMCID: PMCPMC8084877.

22. Organization WH. Global Status Report on Alcohol and Health 2018;(ISBN 978-92-4-156563-9).

23. Kalichman SC, Eaton L. Alcohol-antiretroviral interactive toxicity beliefs as a potential barrier to HIV pre-exposure prophylaxis among men who have sex with men. J Int AIDS Soc. 2017;20(1):21534. doi: 10.7448/IAS.20.1.21534. PubMed PMID: 28715159; PubMed Central PMCID: PMCPMC5577742.

24. Mack N, Evens EM, Tolley EE, Brelsford K, Mackenzie C, Milford C, et al. The importance of choice in the rollout of ARV-based prevention to user groups in Kenya and South Africa: a qualitative study. J Int AIDS Soc. 2014;17(3 Suppl 2):19157. Epub 20140908. doi: 10.7448/IAS.17.3.19157. PubMed PMID: 25224616; PubMed Central PMCID: PMCPMC4164014.

25. Grant RM, Lama JR, Anderson PL, McMahan V, Liu AY, Vargas L, et al. Preexposure chemoprophylaxis for HIV prevention in men who have sex with men. N Engl J Med. 2010;363(27):2587–99. Epub 20101123. doi: 10.1056/NEJMoa1011205. PubMed PMID: 21091279; PubMed Central PMCID: PMCPMC3079639.

26. Chidumwa G, Chimbindi N, Herbst C, Okeselo N, Dreyer J, Zuma T, et al. Isisekelo Sempilo study protocol for the effectiveness of HIV prevention embedded in sexual health with or without peer navigator support (Thetha Nami) to reduce prevalence of transmissible HIV amongst adolescents and young adults in rural KwaZulu-Natal: a 2 x 2 factorial randomised controlled trial. BMC Public Health. 2022;22(1):454. Epub 20220307. doi: 10.1186/s12889-022-12796-8. PubMed PMID: 35255859; PubMed Central PMCID: PMCPMC8900304.

27. Gareta D, Baisley K, Mngomezulu T, Smit T, Khoza T, Nxumalo S, et al. Cohort Profile Update: Africa Centre Demographic Information System (ACDIS) and population-based HIV survey. Int J Epidemiol. 2021;50(1):33–4. Epub 2021/01/14. doi: 10.1093/ije/dyaa264. PubMed PMID: 33437994; PubMed Central PMCID: PMCPMC7938501.

28. Shahmanesh M, Okesola N, Chimbindi N, Zuma T, Mdluli S, Mthiyane N, et al. Thetha Nami: participatory development of a peer-navigator intervention to deliver biosocial HIV prevention for adolescents and youth in rural South Africa. BMC Public Health. 2021;21(1):1393. Epub 20210713. doi: 10.1186/s12889-021-11399-z. PubMed PMID: 34256725; PubMed Central PMCID: PMCPMC8278686.

29. Puryear SB, Balzer LB, Ayieko J, Kwarisiima D, Hahn JA, Charlebois ED, et al. Associations between alcohol use and HIV care cascade outcomes among adults undergoing population-based HIV testing in East Africa. AIDS. 2020;34(3):405–13. doi: 10.1097/QAD.0000000000002427. PubMed PMID: 31725431; PubMed Central PMCID: PMCPMC7046088.

30. Bush K, Kivlahan DR, McDonell MB, Fihn SD, Bradley KA. The AUDIT alcohol consumption questions (AUDIT-C): an effective brief screening test for problem drinking. Ambulatory Care Quality Improvement Project (ACQUIP). Alcohol Use Disorders Identification Test. Arch Intern Med. 1998;158(16):1789–95. doi: 10.1001/archinte.158.16.1789. PubMed PMID: 9738608.

31. Rubinsky AD, Kivlahan DR, Volk RJ, Maynard C, Bradley KA. Estimating risk of alcohol dependence using alcohol screening scores. Drug Alcohol Depend. 2010;108(1-2):29–36. Epub 20091229. doi: 10.1016/j.drugalcdep.2009.11.009. PubMed PMID: 20042299; PubMed Central PMCID: PMCPMC2835806.

32. Africa DoHRoS. 2021 Updated guidelines for the provision of oral pre-exposure prophylaxis (PrEP) to persons at substantial risk of HIV infection 2021:31.

33. Organization WH. Differentiated and simplified pre-exposure prophylaxis for HIV prevention: update to WHO implementation guidance. 2022.

34. Rousseau E, Wu L, Heffron R, Baeten JM, Celum CL, Travill D, et al. Association of sexual relationship power with PrEP persistence and other sexual health outcomes among adolescent and young women in Kenya and South Africa. Front Reprod Health. 2023;5:1073103. Epub 20230530. doi: 10.3389/frph.2023.1073103. PubMed PMID: 37325240; PubMed Central PMCID: PMCPMC10266091.

35. Rao A, Mhlophe H, Comins C, Young K, McIngana M, Lesko C, et al. Persistence on oral pre-exposure prophylaxis (PrEP) among female sex workers in eThekwini, South Africa, 2016-2020. PLoS One. 2022;17(3):e0265434. Epub 20220315. doi: 10.1371/journal.pone.0265434. PubMed PMID: 35290421; PubMed Central PMCID: PMCPMC8923438.

36. Filmer D, Pritchett LH. Estimating wealth effects without expenditure data--or tears: an application to educational enrollments in states of India. Demography. 2001;38(1):115–32. Epub 2001/03/03. doi: 10.1353/dem.2001.0003. PubMed PMID: 11227840.

37. Haberer JE, Baeten JM, Campbell J, Wangisi J, Katabira E, Ronald A, et al. Adherence to antiretroviral prophylaxis for HIV prevention: a substudy cohort within a clinical trial of serodiscordant couples in East Africa. PLoS Med. 2013;10(9):e1001511. Epub 20130910. doi: 10.1371/journal.pmed.1001511. PubMed PMID: 24058300; PubMed Central PMCID: PMCPMC3769210.

38. Marcus JL, Hurley LB, Hare CB, Nguyen DP, Phengrasamy T, Silverberg MJ, et al. Preexposure Prophylaxis for HIV Prevention in a Large Integrated Health Care System: Adherence, Renal Safety, and Discontinuation. J Acquir Immune Defic Syndr. 2016;73(5):540–6. doi: 10.1097/QAI.0000000000001129. PubMed PMID: 27851714; PubMed Central PMCID: PMCPMC5424697.

39. Velloza J, Bacchetti P, Hendrix CW, Murnane P, Hughes JP, Li M, et al. Short- and Long-Term Pharmacologic Measures of HIV Pre-exposure Prophylaxis Use Among High-Risk Men Who Have Sex With Men in HPTN 067/ADAPT. J Acquir Immune Defic Syndr. 2019;82(2):149–58. doi: 10.1097/QAI.0000000000002128. PubMed PMID: 31335588; PubMed Central PMCID: PMCPMC6749964.

40. Miller AP, Shoptaw S, Moucheraud C, Mvududu R, Essack Z, Gorbach PM, et al. Recent Alcohol Use Is Associated With Increased Pre-exposure Prophylaxis (PrEP) Continuation and Adherence Among Pregnant and Postpartum Women in South Africa. J Acquir Immune Defic Syndr. 2023;92(3):204–11. doi: 10.1097/QAI.0000000000003133. PubMed PMID: 36413977; PubMed Central PMCID: PMCPMC9928886.

41. Grov C, Rendina HJ, John SA, Parsons JT. Determining the Roles that Club Drugs, Marijuana, and Heavy Drinking Play in PrEP Medication Adherence Among Gay and Bisexual Men: Implications for Treatment and Research. AIDS Behav. 2019;23(5):1277–86. doi: 10.1007/s10461-018-2309-9. PubMed PMID: 30306433; PubMed Central PMCID: PMCPMC6458096.

42. Sidebottom D, Ekstrom AM, Stromdahl S. A systematic review of adherence to oral pre-exposure prophylaxis for HIV - how can we improve uptake and adherence? BMC Infect Dis. 2018;18(1):581. Epub 20181116. doi: 10.1186/s12879-018-3463-4. PubMed PMID: 30445925; PubMed Central PMCID: PMCPMC6240194.

43. Grammatico MA, Moll AP, Choi K, Springer SA, Shenoi SV. Feasibility of a community-based delivery model for HIV pre-exposure prophylaxis among bar patrons in rural South Africa. J Int AIDS Soc. 2021;24(11):e25848. doi: 10.1002/jia2.25848. PubMed PMID: 34826363; PubMed Central PMCID: PMCPMC8625837.

44. Geldsetzer P, Barnighausen K, Hettema A, McMahon SA, Dalal S, Chase RP, et al. A stepped-wedge randomized trial and qualitative survey of HIV pre-exposure prophylaxis uptake in the Eswatini population. Sci Transl Med. 2020;12(562). doi: 10.1126/scitranslmed.aba4487. PubMed PMID: 32967974; PubMed Central PMCID: PMCPMC8023147.

45. Mudau DO, Mulaudzi FM, Sepeng NV, Anokwuru R. Assessing HIV Pre-exposure Prophylaxis Uptake and Retention Amongst Young Females in Gauteng Province. AIDS Behav. 2023;27(4):1182–7. Epub 20220927. doi: 10.1007/s10461-022-03855-7. PubMed PMID: 36166108.

46. Bekker LG, Giovenco D, Baral S, Dominguez K, Valencia R, Sanchez T, et al. Oral pre-exposure prophylaxis uptake, adherence, and adverse events among South African men who have sex with men and transgender women. South Afr J HIV Med. 2022;23(1):1405. Epub 20221108. doi: 10.4102/sajhivmed.v23i1.1405. PubMed PMID: 36479416; PubMed Central PMCID: PMCPMC9724083.

47. Koss CA, Charlebois ED, Ayieko J, Kwarisiima D, Kabami J, Balzer LB, et al. Uptake, engagement, and adherence to pre-exposure prophylaxis offered after population HIV testing in rural Kenya and Uganda: 72-week interim analysis of observational data from the SEARCH study. Lancet HIV. 2020;7(4):e249–e61. Epub 20200219. doi: 10.1016/S2352-3018(19)30433-3. PubMed PMID: 32087152; PubMed Central PMCID: PMCPMC7208546.

48. Davis CG, Thake J, Vilhena N. Social desirability biases in self-reported alcohol consumption and harms. Addict Behav. 2010;35(4):302–11. Epub 20091110. doi: 10.1016/j.addbeh.2009.11.001. PubMed PMID: 19932936.

49. Latkin CA, Edwards C, Davey-Rothwell MA, Tobin KE. The relationship between social desirability bias and self-reports of health, substance use, and social network factors among urban substance users in Baltimore, Maryland. Addict Behav. 2017;73:133–6. Epub 20170509. doi: 10.1016/j.addbeh.2017.05.005. PubMed PMID: 28511097; PubMed Central PMCID: PMCPMC5519338.

