## Supplemental Material for "Alcohol use and the pre-exposure prophylaxis continuum of care among men in rural South Africa"

**Supporting Information**

**Supplemental Figure 1: Analytic Cohort**

In sensitivity analyses, we included men with 12-month end-line alcohol data. This resulted in 32 men being excluded for missing alcohol data instead of 107 men.

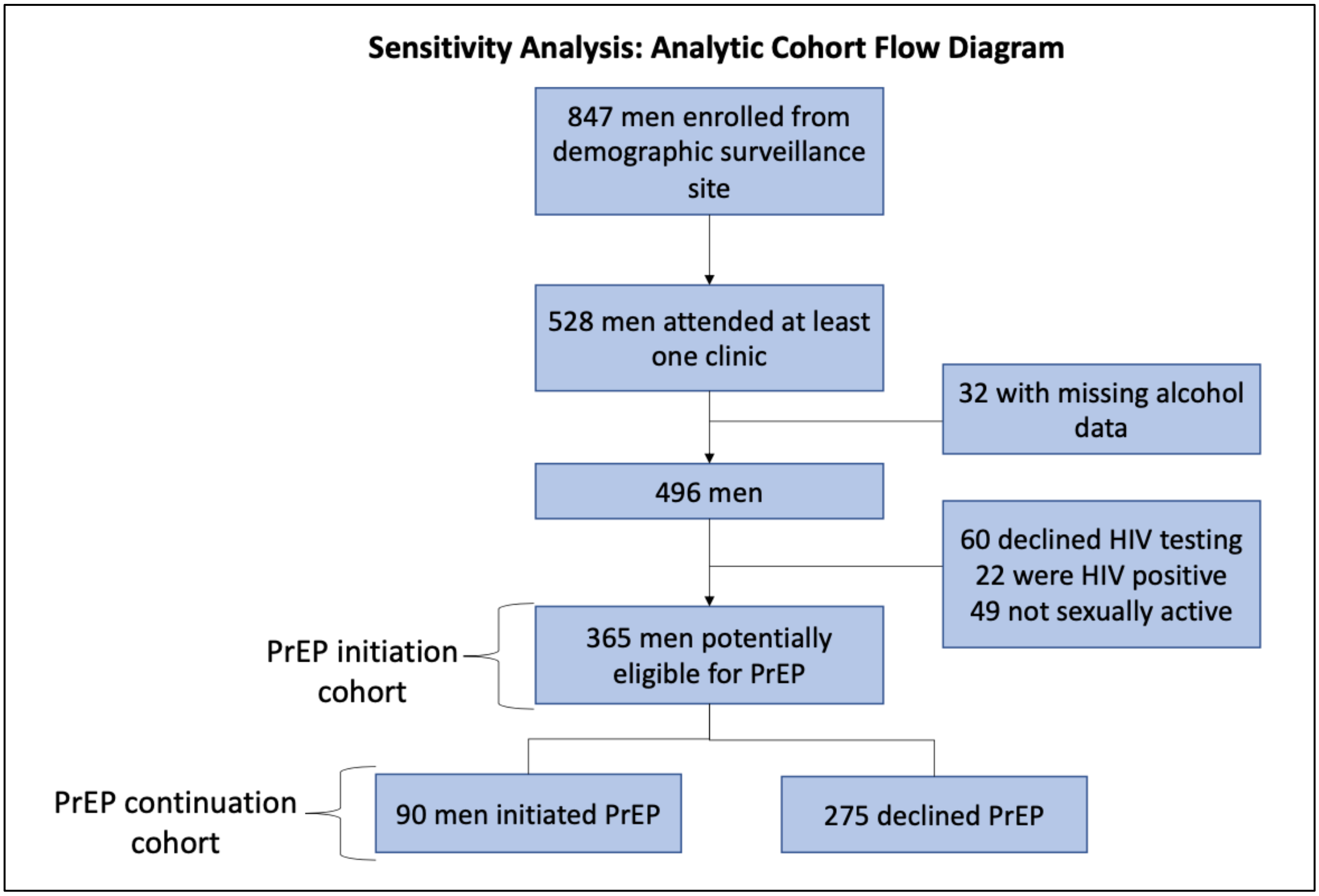

**Supplemental Table 1: Logistic regression models for PrEP initiation and PrEP continuation**

| Characteristic | Univariate Odds Ratio (95%CI) | p-value | Adjusted Odds Ratio (95% CI) | p-value |
|---|---|---|---|---|
| **Model: PrEP initiation (n=365)** | | | | |
| No alcohol | Reference group | | | |
| Low/moderate risk alcohol | 1.91 (0.95-3.85) | 0.071 | 2.08 (0.98-4.43) | 0.056 |
| High/very high risk alcohol | 2.78 (1.59-4.85) | <0.001 | 2.45 (1.31-4.58) | 0.005 |
| Age (years) | 1.07 (1.00-1.14) | 0.039 | 0.99 (0.92-1.07) | 0.801 |
| Socioeconomic status | 0.95 (0.83-1.09) | 0.497 | 0.94 (0.81-1.08) | 0.361 |
| Circumcised | 2.02 (1.34-3.04) | 0.001 | 1.41 (0.89-2.25) | 0.146 |
| Knowledge of sexual partner's HIV status | 2.63 (1.71-4.03) | <0.001 | 1.90 (1.12-1.53) | 0.017 |
| Number of clinic visits | 1.53 (1.29-1.82) | <0.001 | 1.27 (1.05-1.53) | 0.015 |
| **Model: PrEP continuation at 3 months (n=90)** | | | | |
| No alcohol | Reference group | | | |
| Low/moderate risk alcohol | 0.68 (0.14-3.19) | 0.624 | 0.40 (0.07-2.18) | 0.288 |
| High/very high risk alcohol | 1.74 (0.58-5.17) | 0.322 | 0.97 (0.26-3.64) | 0.961 |
| Age (years) | 1.12 (0.98-1.28) | 0.110 | 1.12 (0.94-1.33) | 0.194 |
| Socioeconomic status | 1.17 (0.90-1.51) | 0.242 | 1.16 (0.87-1.55) | 0.32 |
| Circumcised | 3.72 (0.86-15.99) | 0.078 | 5.35 (1.09-26.1) | 0.038 |
| Knowledge of sexual partner's HIV status | 3.73 (1.42-9.78) | 0.007 | 3.08 (1.12-8.50) | 0.029 |
| **Model: PrEP continuation at 6 months (n=90)** | | | | |
| No alcohol | Reference group | | | |
| Low/moderate risk alcohol | 0.70 (0.11-4.32) | 0.701 | 0.47 (0.07-3.26) | 0.445 |
| High/very high risk alcohol | 1.75 (0.50-6.13) | 0.381 | 1.10 (0.25-4.85) | 0.905 |
| Age (years) | 1.10 (0.95-1.28) | 0.204 | 1.07 (0.89-1.29) | 0.448 |
| Socioeconomic status | 1.17 (0.87-1.57) | 0.303 | 1.14 (0.83-1.57) | 0.407 |
| Circumcised | 2.29 (0.54-9.6) | 0.259 | 2.76 (0.59-12.74) | 0.194 |
| Knowledge of sexual partner's HIV status | 3.83 (1.25-11.78) | 0.019 | 3.12 (0.98-9.96) | 0.055 |
